# Novel “SMARTer” Clinical Trial Design Improves Odds of Approval and Can Reduce Study Size By 80%: Modeling Use of a ctDNA “Optimizing Diagnostic” for Early Therapy Switching in Immuno-Oncology Trials

**DOI:** 10.1101/2023.02.20.23286152

**Authors:** Floyd Elliot Taub, Dexiang Gao

## Abstract

Novel pivotal trial designs, that more clearly demonstrate increased benefit over Standard of Care (SOC), especially in oncology and immuno-oncology (IO), are presented. The benefit of therapy is maximized and sample size is dramatically reduced. The novel methodology includes the introduction of a biochemical “Optimizing Diagnostic”, for example a cfDNA test that can detect poor response, when performed early after therapy is begun; this is used to change the therapy of the tested person early in the trial (typically to SOC), before clinical progression. Patients remain in “the new drug first” group, which is compared to SOC. An “Optimizing Diagnostic” is analogous to a “Companion Diagnostic”; both potentially allow approval of drugs that would otherwise fail. A companion diagnostic predicts benefit prior to therapy, the optimizing diagnostic (more accurately) predicts likelihood of benefit after initial therapy. Those patients deemed less likely to respond remain in the novel drug first arm, but are switched to SOC. A Sequential Multiple Assignment Randomized Trial” (SMART) design is proposed to evaluate if switching to SOC or SOC plus continuing the novel therapy is most beneficial. These designs will allow approval of therapy paths including novel agents when the novel agent could not be approved without this design. A good optimizing test may reduce the number of patients needed by 80%, dramatically reducing cost and time; more patients benefit and accrual is easier.

**Key Points:**

- A new clinical trial design focused on testing a path that begins with a novel regimen (IO is featured) is presented.
- The path includes an “optimizing diagnostic” that determines, early during treatment, if a patient should remain on the new regimen.
- Companion diagnostics define the path at a pre-treatment stage, optimizing diagnostics define the path early during treatment. Changes in therapy, typically to the SOC, is made based on the post-test probability of success.
- The novel path is significantly more likely to lead to approval than the novel regimen alone.
- Use of the novel method can reduce the size of a trial by 80%, and allow approval of the path, when approval of the novel regimen, based on a head-to-head trial vs SOC, would not be possible.

## Introduction

The classic randomized clinical trial (RCT) has evolved since Johannes Fibiger, in 1898^1^, designed a trial to determine the best treatment for future patients with acute diphtheria by comparing standard of care (SOC) to SOC plus anti-serum. Today it would be critiqued as both quasi-randomized and unblinded^2^. However, to this day the principal stands: a new therapy is compared to the current SOC. The results are used to both seek regulatory approval of a new drug and provide convincing data supporting its use in future patients. This paper will focus on a novel designs to seek approval and chose therapies appropriate for many chronic diseases and especially for cancer.

The gold standard RCT has evolved into the double-blind, randomized controlled trial (DBRCT). A series of DBRCTs superbly determine which therapy is better in the population in the trial. In the situation of acute disease, it identifies the therapy to be chosen. However, in first-tier healthcare nations, death is overwhelmingly due to chronic disease. In clinical practice patients typically are given multiple drugs in sequence, or in combination. Neither the classic RCT or DBRCT typically used for approval, addresses this real-world medical practice of multiple sequential therapies. Murphy, in 2005 proposed Sequential Multiple Assignment

Randomized (SMAR) trials for behavior modification, but noted they were best used for hypothesis generation about sequential treatment and had “insurmountable” high dimensionality issues. In 2007 Collins^3^ proposed an eHealth “Sequential Multiple Assignment Randomized Trial” (SMART) to experimentally test what sequence of eHealth therapies are best. Cardio-vascular disease (CVD) is the most frequent cause of death in many countries and a series of sequential therapies to reduce risk factors and fatal events is SOC. In 2022 Lambert^4^ applied formal SMART design to evaluate web-based stress management in CVD. Current prevention practice typically mandates starting with one drug (often a statin), prompt *<u>biochemically testing</u>* of effectiveness to determine if it reduces cholesterol-based risk factors adequately, and adjusting therapy, which often includes higher doses and/or additional drugs to achieve the desired (biochemical) result.

**Cancer is the second leading cause of death; again, this is often a chronic disease. While some patients are cured initially, almost all of the 600**,**000 US patients who die of cancer have had sequential therapy**. As in CVD, clinical decision trees have evolved for cancer, but strikingly, except for testing no continuing therapy vs maintenance therapy^5,6^, the first discussion of a formal SMART design for oncology was in 2018^7^ when Kidwell, et al., proposed an immuno-oncology (IO) trial that, consistent with Murphy’s original concern of high dimensionality, resulted in 7 to 10 comparisons. This had limited practicality and, as the authors noted, led to statistical complexity; it has not been executed.

Herein we propose the first practical ***SMART trial design to enhance registration* (SMARTer)** of a novel therapy path. It is the first design to use a biochemical “Optimizing Diagnostic”, measured before progressive disease, to revise therapy to include SOC. In Section A below, the design specifies changing therapy to SOC; in Section B, it specifies a second randomization to either SOC or SOC plus new drug(s). In all cases patients first treated with the new drug remain within the experimental arm for statistical analysis. Both these designs result in capturing within the experimental group some benefit of the new therapy and the SOC.

**The combined, for some patients sequential, benefit is compared to SOC**. Modeling shows this dramatically reduces the patients needed to prove benefit of a new therapy. Further it will identify benefit of a therapy path including the new therapy, when benefit of the new drug without this novel therapy path, is not present. Thus, it decreases the investment (patients, time and dollars) to approval for some therapies and can detect approvable response when none would be detected with standard trial designs.

## Section A: Introduction of an Optimizing Diagnostic without Sequential Randomization

### Trial Design Description

IO drugs have revolutionized oncology as they may cure even late-stage patients and are used as an example herein. While indications and situations vary greatly, they often give durable responses in only 10% of the population, even when appropriate patients are preselected with a predictive companion diagnostic. Long-term benefit varies from 0 to 100% depending on the type of cancer and clinical situation; in many situations about ∼20-50% show some benefit. Unfortunately, again depending on the situation, about, 15-20% may have more than doubling in the growth rate (Hyperprogressive Disease or HPD) of their cancer^8, 9, 10^, both Russo et al^11^, and Yildirim et al.^12^ report NSCLC patients have ∼30% incidence of HPD.

DNA methods, including cfDNA, have become SOC for detecting actionable mutations. They can also be used soon after therapy is started to monitor drug effectiveness. Herein we propose using such information to improve clinical trial as well as patient outcomes. cfDNA testing soon after therapy begins can allow much greater accuracy in determining if a patient will respond. While currently expensive and time consuming, there are many reports that increasing circulating tumor DNA, ctDNA, accurately predicts if therapy is doomed to fail. While cfDNA is not itself an approvable endpoint, the FDA allows, even encourages^13^, ctDNA levels being used in a clinical trial.

Figure 1 illustrates a clinical trial path from left to right as time progresses. It starts with a companion diagnostic “Tailoring test” followed by classic randomization to SOC vs. a new IO treatment first. Patients always remain in these originally assigned groups; the statistical plan compares these groups. On the day of therapy initiation, before therapy, a quantitative ctDNA determination is performed. (Baseline imaging tests historically precede treatment by significantly longer and thus may provide a less accurate baseline.) During the initial therapy the quantitative ctDNA is repeated; it is designated the Optimizing Diagnostic. For convenience, this may be just prior to each IO infusion, which are typically 3 weeks apart. The likelihood of a patient responding is reassessed. If optimizing diagnostic testing predicts likely IO failure, that person is transferred to SOC but remains in the “IO First” group. While the companion diagnostic test may have selected a group anticipated to have, for example a 50% chance of response, (and SOC was anticipated to have a 30% chance of response) the on therapy optimizing diagnostic test might indicate the person now has only a 0-10% chance of response. Thus, this person will likely do better (and deserves) SOC, as some benefit, maybe still 30%, from SOC is anticipated. Exact models and numbers are presented below. Each New IO therapy first patient, and this trial group as a whole, benefits from this increasingly personalized care. They may experience significant benefit from the new IO and from SOC; the sum is greater than either alone. The target is that those patients who will be cured or improved with IO will achieve that benefit, while the IO hyper-progressors and progressors will get the benefit of SOC. The below analysis models if, and how much, the sum of IO plus sequential SOC, is better than SOC.

**Figure 1.**
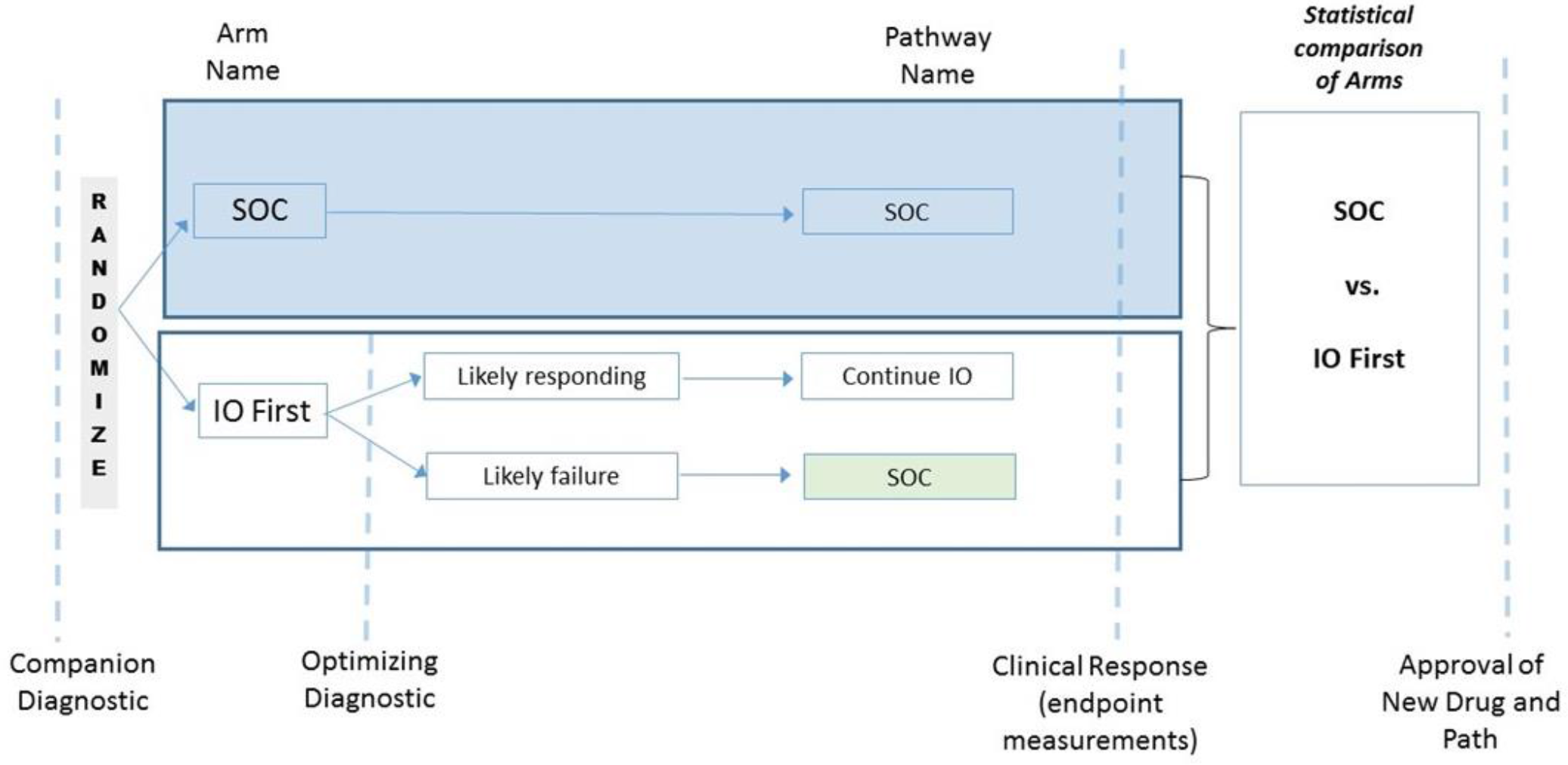
Introduction of an Optimizing Diagnostic

The benefit depends on the accuracy of the optimizing diagnostic test. Various levels of accuracy of the optimizing test are modeled in the examples below. **The sensitivity is defined as the proportion of patients who test positive for likely to progress, among the true disease progressive subjects; the specificity is defined as the proportion of test negative subjects among the true non-progressive subjects**. Various levels of sensitivity and specificity are presented in the below examples.

**<u>Modeling Scenario 1</u>: Benefit resulting from early treatment switching from IO to SOC for subjects found to be at high risk of progressing on IO based on an early optimizing diagnostic**

Within this scenario #1, we consider a situation in which IO is somewhat better than SOC as a single agents, two assumptions regarding potential IO effect on later SOC: in 1.1) the effectiveness of subsequent SOC is presumed to be the same as if the SOC was given initially; in 1.2) SOC effectiveness was presumed to decrease if given after IO (potentially due to delayed treatment). In scenario 2 we consider both the above scenarios but assume that as single agents the IO and SOC have the same performance. Based on literature reviewed in the discussion, there is a possibility that IO sensitizes to subsequent SOC, this is modeled in two scenarios: 3.1 models a situation when the IO is better than SOC as single agents, 3.2 models a situation in which they are equal as single agents.

In these examples it is assumed that the mechanism of action of the two drugs is distinct and cross resistance is not/or minimally present. This criterion is met for comparing IO to SOC chemotherapy or growth pathway targeted therapy.

One likely scenario is that the IO is more effective than SOC, however the difference is small; this is assumed in Scenario 1. Table 1.1 examines a scenario in which SOC is assumed to have an effectiveness of 30% (regardless of when administered), while IO is assumed to be 40% effective.

**Table 1.1.** Benefit of switching early after using “Optimizing Diagnostic” test in the IO First arm with the scenario IO response rate of 40% and SOC response rate of 30%; SOC effectiveness remains the same after IO.

| Sensitivity | Specificity | PPV | NPV | % Total benefit* | % Benefit over SOC** |
| --- | --- | --- | --- | --- | --- |
| 0.95 | 0.95 | 92.68 | 96.61 | 55.7 | 85.7 |
| 0.95 | 0.90 | 86.36 | 96.43 | 54.8 | 82.7 |
| 0.95 | 0.80 | 76.00 | 96.00 | 53.0 | 76.7 |
| 0.95 | 0.70 | 67.86 | 95.45 | 51.2 | 70.7 |
| 0.90 | 0.95 | 92.31 | 93.44 | 54.3 | 81.0 |
| 0.90 | 0.90 | 85.71 | 93.10 | 53.4 | 78.0 |
| 0.90 | 0.80 | 75.00 | 92.31 | 51.6 | 72.0 |
| 0.90 | 0.70 | 66.67 | 91.30 | 49.8 | 66.0 |
| 0.80 | 0.95 | 91.43 | 87.69 | 51.5 | 71.7 |
| 0.80 | 0.90 | 84.21 | 87.10 | 50.6 | 68.7 |
| 0.80 | 0.80 | 72.73 | 85.71 | 48.8 | 62.7 |
| 0.80 | 0.70 | 64.00 | 84.00 | 47.0 | 56.7 |
| 0.70 | 0.95 | 90.32 | 82.61 | 48.7 | 62.3 |
| 0.70 | 0.90 | 82.35 | 81.82 | 47.8 | 59.3 |
| 0.70 | 0.80 | 70.00 | 80.00 | 46.0 | 53.3 |
| 0.70 | 0.70 | 60.87 | 77.78 | 44.2 | 47.3 |

\*Total benefit = 100 \*((percentage of test positive[response to IO]) \* PPV + (percentage of test negative [no response to IO]) \* SOC later).
\*\* Benefit over SOC = 100 \* (Total Benefit – SOC) / SOC). Values are %.

The top row indicates that using sequential treatment based on an optimizing diagnostic test with a 95% sensitivity and specificity (95/95), the improvement in patient outcome an [IO first, test, continue treatment with IO or SOC as indicated] scenario compared to an [IO only] scenario would be 55.7%. Simple testing and using the result to decide if IO should be continued or if SOC is better for that patient improves the outcome in the experimental arm 55.7%. This would be a striking 85.67% increase over SOC, this contrasts with the classic head-to-head comparison (no optimizing test or switching) in which the benefit would be only 33%, a 2.6x improvement. Even a poor optimizing diagnostic test with sensitivity and specificity of only 70% (70/70) results in an “IO First” arm response rate of 44.2% and results in a 47.3% improvement over SOC.

Figure 2 shows these results graphically. Dependence on both sensitivity and specificity are illustrated. Not surprisingly, higher levels of both increase the benefit of using an optimizing diagnostic test.

**Figure 2.**
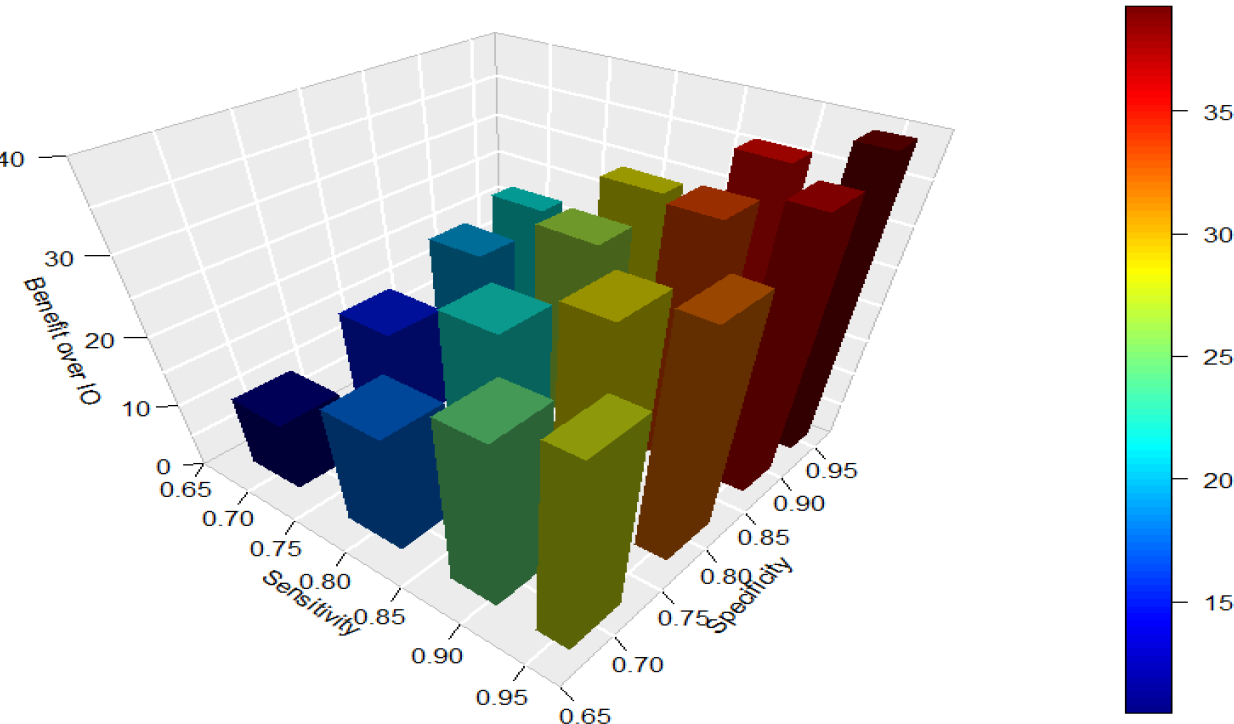

One implication of the improvement in outcome with a good optimizing diagnostic and sequential therapy is that fewer patients are required to see a statistically significant benefit. Table 1.2 is a power calculation for the above scenario.

**Table 1.2.** Benefit of switching early after using “Optimizing Diagnostic” test in the IO First arm with the scenario IO response rate of 40%, SOC response rate of 30%, and SOC response rate 20% if given later.

| Sensitivity | Specificity | PPV | NPV | % Total benefit* | % Benefit over SOC** |
| --- | --- | --- | --- | --- | --- |
| 0.95 | 0.95 | 92.68 | 96.61 | 49.8 | 66.0 |
| 0.95 | 0.90 | 86.36 | 96.43 | 49.2 | 64.0 |
| 0.95 | 0.80 | 76.00 | 96.00 | 48.0 | 60.0 |
| 0.95 | 0.70 | 67.86 | 95.45 | 46.8 | 56.0 |
| 0.90 | 0.95 | 92.31 | 93.44 | 48.2 | 60.7 |
| 0.90 | 0.90 | 85.71 | 93.10 | 47.6 | 58.7 |
| 0.90 | 0.80 | 75.00 | 92.31 | 46.4 | 54.7 |
| 0.90 | 0.70 | 66.67 | 91.30 | 45.2 | 50.7 |
| 0.80 | 0.95 | 91.43 | 87.69 | 45.0 | 50.0 |
| 0.80 | 0.90 | 84.21 | 87.10 | 44.4 | 48.0 |
| 0.80 | 0.80 | 72.73 | 85.71 | 43.2 | 44.0 |
| 0.80 | 0.70 | 64.00 | 84.00 | 42.0 | 40.0 |
| 0.70 | 0.95 | 90.32 | 82.61 | 41.8 | 39.3 |
| 0.70 | 0.90 | 82.35 | 81.82 | 41.2 | 37.3 |
| 0.70 | 0.80 | 70.00 | 80.00 | 40.0 | 33.3 |
| 0.70 | 0.70 | 60.87 | 77.78 | 38.8 | 29.3 |

\*Total benefit = 100 \*((percentage of test positive[response to IO]) \* PPV + (percentage of test negative [no response to IO]) \* SOC later)
\*\* Benefit over SOC = 100 \* (Total Benefit – SOC) / SOC)

**Without an optimizing diagnostic and rapid switching, 355 patients** in each arm (710 total) are needed to test the 10% points difference between SOC and the original IO (no switching) with a 80% power using a two-sided test.

**Table 1.1a.** Sample Size calculation for case 1.1, with 80% power, 2-sided test with alpha of 0.05, assuming a 10% difference in benefit.

| Sensitivity | Specificity | PPV | NPV | Total benefit | Sample Size | % Reduction in Sample Size |
| --- | --- | --- | --- | --- | --- | --- |
| 0.95 | 0.95 | 92.68 | 96.61 | 55.7 | 57 | **84** |
| 0.95 | 0.90 | 86.36 | 96.43 | 54.8 | 61 | **83** |
| 0.95 | 0.80 | 76.00 | 96.00 | 53.0 | 71 | **80** |
| 0.95 | 0.70 | 67.86 | 95.45 | 51.2 | 83 | **77** |
| 0.90 | 0.95 | 92.31 | 93.44 | 54.3 | 63 | **82** |
| 0.90 | 0.90 | 85.71 | 93.10 | 53.4 | 68 | **81** |
| 0.90 | 0.80 | 75.00 | 92.31 | 51.6 | 80 | **77** |
| 0.90 | 0.70 | 66.67 | 91.30 | 49.8 | 95 | **73** |
| 0.80 | 0.95 | 91.43 | 87.69 | 51.5 | 81 | **77** |
| 0.80 | 0.90 | 84.21 | 87.10 | 50.6 | 88 | **75** |
| 0.80 | 0.80 | 72.73 | 85.71 | 48.8 | 105 | **70** |
| 0.80 | 0.70 | 64.00 | 84.00 | 47.0 | 127 | **64** |
| 0.70 | 0.95 | 90.32 | 82.61 | 48.7 | 106 | **70** |
| 0.70 | 0.90 | 82.35 | 81.82 | 47.8 | 116 | **67** |
| 0.70 | 0.80 | 70.00 | 80.00 | 46.0 | 143 | **60** |
| 0.70 | 0.70 | 60.87 | 77.78 | 44.2 | 180 | **49** |

Sample size in each arm required to test the difference between SOC and the Total benefit in the IO arm resulting from switching IO non-responders to SOC, assuming equal sample size between the two arms. R package 'pwr' is used for the power calculation.

**Strikingly less than 1/5 the number of patients is required; the sample size required falls from 355 to 57 in each arm, (710 vs. 114 total, 84% reduction) using a good optimizing test (95/95)**. Even with a poor test (70/70) the number of patients needed is reduced from 355 in each arm to 180.

We next examined a scenario (designated **Scenario 1.2)** in which it was assumed that **SOC had a 33% decrease in effectiveness if delayed** (30% if given first, 20% if given later), to see if this negated the benefit of testing and sequential therapy in the “IO First” treatment arm.

While, as expected, the benefit of an optimizing diagnostic and rapid switching was lower than in scenario 1.1, with a test having a sensitivity and specificity of 95% there was still a 49.8% response rate in the sequential therapy group which is a surprisingly large 66% benefit over SOC. If the test was poor, (70/80 or less) and SOC lost effectiveness if delayed, there was still a benefit over SOC, but it was not better than IO alone in this scenario where IO alone was 30% better than SOC.

The sample size (SS) calculation for this example 1.2 is given below. A sample size of 355 in each arm, for the standard no optimizing test design, is needed; in contrast, only 95 patients per arm would be needed with a good optimizing test. This is a 73% savings.

**Table 1.2a.** Sample size calculation for case 1.2, with 80% power, 2-sided test with alpha of 0.05, assuming a 10% difference in benefit.

| Sensitivity | Specificity | PPV | NPV | Total benefit | Sample Size | % Reduction in Sample Size |
| --- | --- | --- | --- | --- | --- | --- |
| 0.95 | 0.95 | 92.68 | 96.61 | 49.8 | 95 | 73 |
| 0.95 | 0.90 | 86.36 | 96.43 | 49.2 | 100 | 72 |
| 0.95 | 0.80 | 76.00 | 96.00 | 48.0 | 114 | 68 |
| 0.95 | 0.70 | 67.86 | 95.45 | 46.8 | 130 | 63 |
| 0.90 | 0.95 | 92.31 | 93.44 | 48.2 | 111 | 69 |
| 0.90 | 0.90 | 85.71 | 93.10 | 47.6 | 119 | 66 |
| 0.90 | 0.80 | 75.00 | 92.31 | 46.4 | 139 | 61 |
| 0.90 | 0.70 | 66.67 | 91.30 | 45.2 | 158 | 55 |
| 0.80 | 0.95 | 91.43 | 87.69 | 45.0 | 162 | 54 |
| 0.80 | 0.90 | 84.21 | 87.10 | 44.4 | 175 | 51 |
| 0.80 | 0.80 | 72.73 | 85.71 | 43.2 | 207 | 42 |
| 0.80 | 0.70 | 64.00 | 84.00 | 42.0 | 250 | 30 |
| 0.70 | 0.95 | 90.32 | 82.61 | 41.8 | 258 | 27 |
| 0.70 | 0.90 | 82.35 | 81.82 | 41.2 | 285 | 20 |
| 0.70 | 0.80 | 70.00 | 80.00 | 40.0 | 355 | 0 |

Sample size in each arm required to test the difference between the SOC and the Total Benefit in the IO arm resulting from switching IO non-responders to SOC, assuming equal sample size between the two arms. R package “pwr” is used for the power calculation.

**We next asked if this method could result in approval of a sequence starting with the new IO, even if the IO alone was no more effective than SOC alone**. Example 2.1 models a situation in which both the IO and SOC have a 30% effectiveness. Under this situation, without this trial design, approval would be unlikely without a major difference in side effects, convenience, etc. The first assumption in this section (2.1) is that SOC effectiveness does not change due to the delay in administering it.

**Table 2.1.** Benefit of switching early after using “Optimizing Diagnostic” test in the IO First arm with the scenario IO and SOC equal as single agents (30%) and the SOC effectiveness remains the same (30%) if given later, after IO.

| Sensitivity | Specificity | PPV | NPV | Total benefit* | Benefit over SOC** |
| --- | --- | --- | --- | --- | --- |
| 0.95 | 0.95 | 89.06 | 97.79 | 48.9 | **63.0** |
| 0.95 | 0.90 | 80.28 | 97.67 | 47.9 | 59.5 |
| 0.95 | 0.80 | 67.06 | 97.39 | 45.8 | 52.5 |
| 0.95 | 0.70 | 57.58 | 97.03 | 43.7 | 45.5 |
| 0.90 | 0.95 | 88.52 | 95.68 | 47.9 | 59.5 |
| 0.90 | 0.90 | 79.41 | 95.45 | 46.8 | 56.0 |
| 0.90 | 0.80 | 65.85 | 94.92 | 44.7 | 49.0 |
| 0.90 | 0.70 | 56.25 | 94.23 | 42.6 | 42.0 |
| 0.80 | 0.95 | 87.27 | 91.72 | 45.8 | 52.5 |
| 0.80 | 0.90 | 77.42 | 91.30 | 44.7 | 49.0 |
| 0.80 | 0.80 | 63.16 | 90.32 | 42.6 | 42.0 |
| 0.80 | 0.70 | 53.33 | 89.09 | 40.5 | 35.0 |
| 0.70 | 0.95 | 85.71 | 88.08 | 43.7 | 45.5 |
| 0.70 | 0.90 | 75.00 | 87.50 | 42.6 | 42.0 |
| 0.70 | 0.80 | 60.00 | 86.15 | 40.5 | 35.0 |
| 0.70 | 0.70 | 50.00 | 84.48 | 38.4 | 28.0 |

\*Total benefit = 100 \* ((percentage of test positive[response to IO]) \* PPV + (percentage of test negative [no response to IO]) \* SOC later)
\*\* Benefit over SOC = 100 \* (Total Benefit – SOC) / SOC

**The top row indicates that an optimizing diagnostic with a 95% sensitivity and specificity would improve patient outcome to 48.9% in the experimental group (IO first) and thus this sequential treatment would have a 63% improvement of SOC**. This level of improvement is likely significant for patient care and approval. Even a poor test with sensitivity and specificity of only 70% improves new arm outcome by 43.7% and results in a 45.5% improvement over SOC, suggesting approval of this sequential path.

We next asked if this remarkable approval of a sequential path, when the first drug alone could not be approved, was still true if the effectiveness of SOC decreased if it was delayed. Table 2.2 models this.

**Table 2.2.** Benefit of switching early after using “Optimizing Diagnostic” test in the IO First arm with the scenario IO and SOC equal as single agents and the SOC effectiveness decreases after IO. SOC=IO=30%, SOC=20% if given later.

| Sensitivity | Specificity | PPV | NPV | Total benefit | Benefit over SOC |
| --- | --- | --- | --- | --- | --- |
| 0.95 | 0.95 | 89.06 | 97.79 | 42.1 | 40.3 |
| 0.95 | 0.90 | 80.28 | 97.67 | 41.4 | 38.0 |
| 0.95 | 0.80 | 67.06 | 97.39 | 40.0 | 33.3 |
| 0.95 | 0.70 | 57.58 | 97.03 | 38.6 | 28.7 |
| 0.90 | 0.95 | 88.52 | 95.68 | 49.0 | 36.3 |
| 0.90 | 0.90 | 79.41 | 95.45 | 42.0 | 34.0 |
| 0.90 | 0.80 | 65.85 | 94.92 | 38.8 | 29.3 |
| 0.90 | 0.70 | 56.25 | 94.23 | 37.4 | 24.7 |
| 0.80 | 0.95 | 87.27 | 91.72 | 38.5 | 28.3 |
| 0.80 | 0.90 | 77.42 | 91.30 | 37.8 | 26.0 |
| 0.80 | 0.80 | 63.16 | 90.32 | 36.4 | 21.3 |
| 0.80 | 0.70 | 53.33 | 89.09 | 35.0 | 16.7 |
| 0.70 | 0.95 | 85.71 | 88.08 | 36.1 | 20.3 |
| 0.70 | 0.90 | 75.00 | 87.50 | 35.4 | 18.0 |
| 0.70 | 0.80 | 60.00 | 86.15 | 34.0 | 13.3 |
| 0.70 | 0.70 | 50.00 | 84.48 | 32.6 | 8.7 |
| *Total benefit = 100 * ((percentage of test positive [response to IO]) * PPV + (percentage of test negative [no response to IO]) * SOC later) |  |  |  |  |  |
| ** Benefit over SOC = 100 * (Total Benefit – SOC) / SOC |  |  |  |  |  |

While the benefit of switching was less, with a test having a sensitivity and specificity of 95% there was still a 42.1% response rate in the sequential therapy group and a 40% benefit over SOC. While these numbers would significantly improve patient care and chance of approval, if the test was poor, 70% sensitivity and specificity, the benefit over SOC fell to only 8.67%. Thus, even if the new drug alone is un-approvable alone, as it has not benefit over SOC, with a good, or even moderately good optimizing diagnostic test, sequential therapy based on an optimizing diagnostic shows apparently approvable benefit over SOC.

While intuition suggests a delay in SOC would result in some decrease in benefit, as reviewed in the discussion section, a growing body of evidence suggests IO potentiates SOC in certain situations, even when the IO alone is not effective, thus we modeled this possibility. Below (Table 3) we model the benefit of sequential therapy if IO sensitizes to subsequent SOC.

**Table 3.1.**
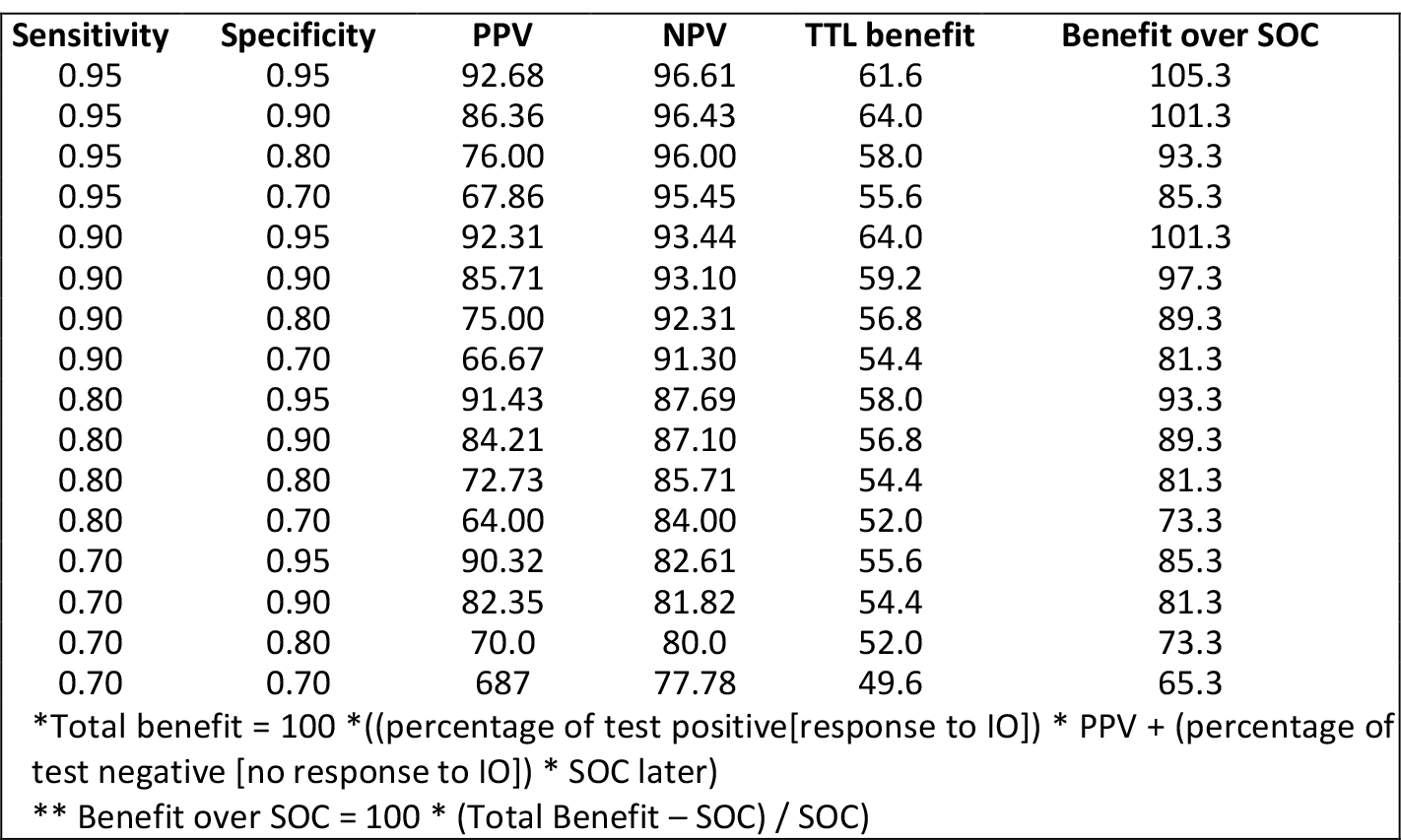
Benefit of switching early after using “Optimizing Diagnostic” test in the IO First arm with the scenario IO is better than SOC and sensitizes to SOC. SOC=30%, IO=40%, SOC=40% if given later.

| Sensitivity | Specificity | PPV | NPV | TTL benefit | Benefit over SOC |
| --- | --- | --- | --- | --- | --- |
| 0.95 | 0.95 | 92.68 | 96.61 | 61.6 | 105.3 |
| 0.95 | 0.90 | 86.36 | 96.43 | 64.0 | 101.3 |
| 0.95 | 0.80 | 76.00 | 96.00 | 58.0 | 93.3 |
| 0.95 | 0.70 | 67.86 | 95.45 | 55.6 | 85.3 |
| 0.90 | 0.95 | 92.31 | 93.44 | 64.0 | 101.3 |
| 0.90 | 0.90 | 85.71 | 93.10 | 59.2 | 97.3 |
| 0.90 | 0.80 | 75.00 | 92.31 | 56.8 | 89.3 |
| 0.90 | 0.70 | 66.67 | 91.30 | 54.4 | 81.3 |
| 0.80 | 0.95 | 91.43 | 87.69 | 58.0 | 93.3 |
| 0.80 | 0.90 | 84.21 | 87.10 | 56.8 | 89.3 |
| 0.80 | 0.80 | 72.73 | 85.71 | 54.4 | 81.3 |
| 0.80 | 0.70 | 64.00 | 84.00 | 52.0 | 73.3 |
| 0.70 | 0.95 | 90.32 | 82.61 | 55.6 | 85.3 |
| 0.70 | 0.90 | 82.35 | 81.82 | 54.4 | 81.3 |
| 0.70 | 0.80 | 70.0 | 80.0 | 52.0 | 73.3 |
| 0.70 | 0.70 | 68.7 | 77.78 | 49.6 | 65.3 |

\*Total benefit = 100 \* ((percentage of test positive[response to IO]) \* PPV + (percentage of test negative [no response to IO]) \* SOC later)
\*\* Benefit over SOC = 100 \* (Total Benefit – SOC) / SOC)

Strikingly potentiation of SOC from 30% to 40% effective, in the setting of IO being 40% effective while SOC is 30% effective, results in 105% benefit over SOC with a good optimizing diagnostic test (95/95).

Lastly, we asked if an IO was no better than SOC but potentiated subsequent SOC from 30% to 40% would sequential therapy yield improvement significant for clinical use and potentially approval.

**Table 3.2.** Benefit of switching early after using “Optimizing Diagnostic” test in the IO First arm with the scenario IO is equal to SOC and sensitizes to SOC. SOC=IO=30%, SOC=40% if given later.

| Sensitivity | Specificity | PPV | NPV | Total benefit | Benefit over SOC |
| --- | --- | --- | --- | --- | --- |
| 0.95 | 0.95 | 89.06 | 97.79 | 55.7 | 85.7 |
| 0.95 | 0.90 | 80.28 | 97.67 | 54.3 | 81.0 |
| 0.95 | 0.80 | 67.06 | 97.39 | 51.5 | 71.7 |
| 0.95 | 0.70 | 57.58 | 97.03 | 48.7 | 62.3 |
| 0.90 | 0.95 | 88.52 | 95.68 | 54.8 | 82.7 |
| 0.90 | 0.90 | 79.41 | 95.45 | 53.4 | 78.0 |
| 0.90 | 0.80 | 65.85 | 94.92 | 50.6 | 68.7 |
| 0.90 | 0.70 | 56.25 | 94.23 | 47.8 | 59.3 |
| 0.80 | 0.95 | 87.27 | 91.72 | 53.0 | 76.7 |
| 0.80 | 0.90 | 77.42 | 91.30 | 51.6 | 72.0 |
| 0.80 | 0.80 | 63.16 | 90.32 | 48.8 | 62.7 |
| 0.80 | 0.70 | 53.33 | 89.09 | 46.0 | 53.3 |
| 0.70 | 0.95 | 85.71 | 88.08 | 51.2 | 70.7 |
| 0.70 | 0.90 | 75.00 | 87.50 | 49.8 | 66.0 |
| 0.70 | 0.80 | 60.00 | 86.15 | 47.0 | 56.7 |
| 0.70 | 0.70 | 50.0 | 84.48 | 44.2 | 47.3 |

\*Total benefit = 100 \* ((percentage of test positive[response to IO]) \* PPV + (percentage of test negative [no response to IO]) \* SOC later)
\*\* Benefit over SOC = 100 \* (Total Benefit – SOC) / SOC)

In this situation of IO being no better than SOC alone, but showing a potentiation of SOC from 30% to 40% effective, a 56% response and 86% improvement over SOC is seen with sequential therapy. Thus, we find another striking example of an un-approvable drug being potentially approvable if used to initiate a sequence and followed with an optimizing diagnostic.

## Section B: Sequential Randomization in Addition to an Optimizing Diagnostic

With a IO success rate of 30%, many patients will be transferred to SOC. This presents an opportunity for a subsequent randomization to answer the critical question of whether SOC alone, or SOC with simultaneous IO is better in this IO alone resistant subgroup. As there can be classic randomization of this subgroup, and to the degree side effects don’t prevent it, blinding, the question of which is better can be answered. (Because it is a select sub-group, who may have more resistant disease, the outcome in these subgroups cannot be compared to either SOC alone or IO alone.) However, it is trivial to extrapolate what would have happened if everyone the optimizing diagnostic suggested would be unlikely to do well on IO alone had transferred to the more optimum therapy (SOC alone or SOC plus IO); using this extrapolated result, this optimum path for those initially started on IO can be compared with the SOC.

The trial flow is shown in Figure 3; it adds to that shown in Figure 1 a second randomization (in red) for those patients likely to fail IO alone based on the optimizing diagnostic.

**Figure 3.**
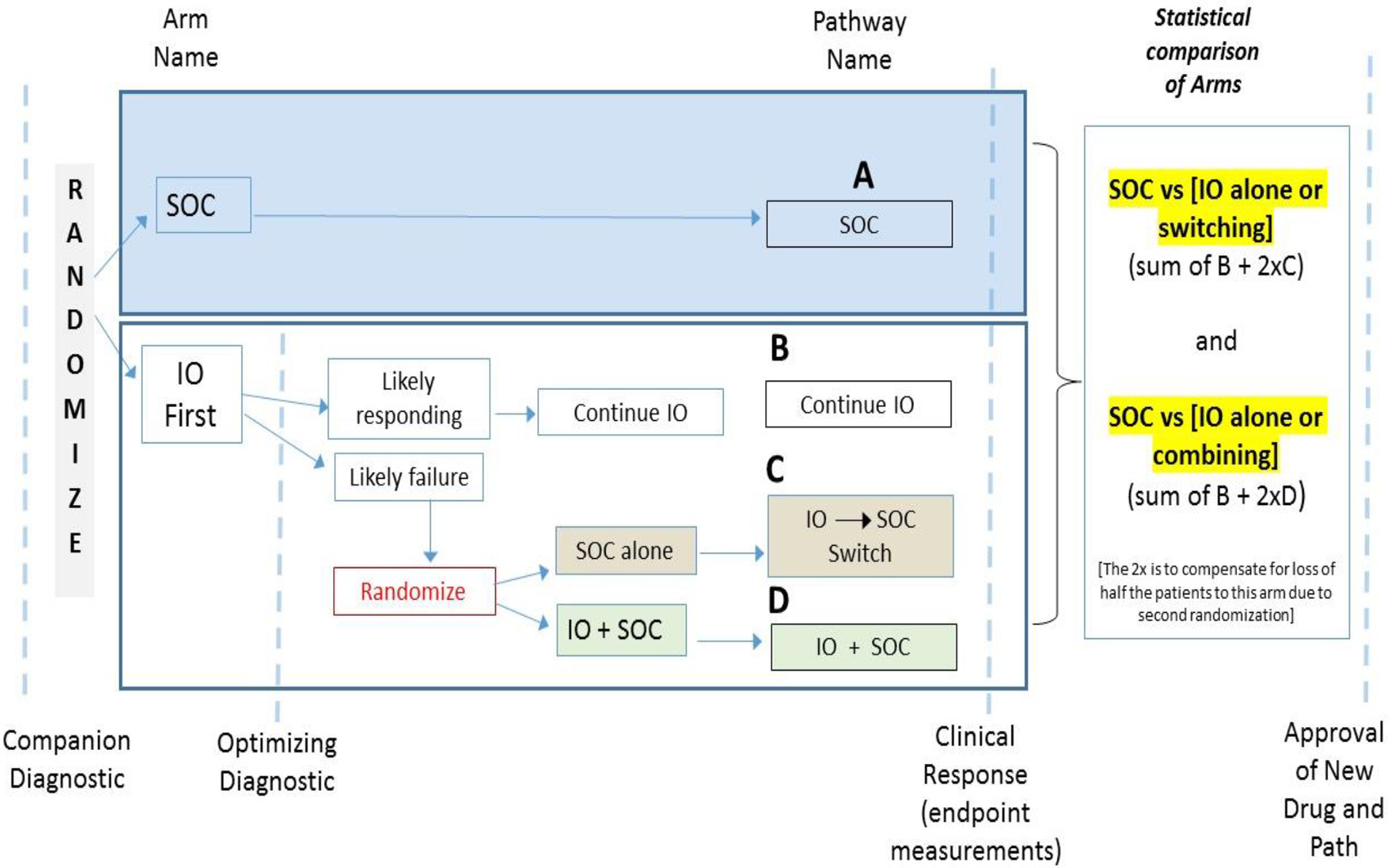
Optimizing diagnostic with sequential randomization

When the optimizing diagnostic test indicates likely failure of IO alone (in many trials the most likely outcome) that patient is randomized to either switching to SOC, which has benefits as illustrated in section 1 above, or adding SOC while continuing IO. This sequential randomization (the above examples show sequential treatment based on the optimizing test but not a second randomization) allows direct comparison of benefit and side effects of 1) SOC alone and 2) SOC plus IO, in a group resistant to IO alone. From the outcome the best of these two can be chosen and calculations comparing the benefit of the optimal path if the total group that started on IO were to have had the optimal path for them. The numbers in the above table are also applicable to this analysis. The exact nature of the second therapy does not change the mathematical analysis.

## Discussion

In situations of acute fatal disease, the scientific question of which drug is “better” and the medical question of which drug is best to administer, are the same. In contrast, for nonfatal chronic illness, physicians often start with the least toxic (medically and/or financially) and only proceed to the “more expensive/bigger gun” alternatives as needed. In many cases a sequence of drugs or “path” is used to achieve the best balance of toxicity and benefit. Historically, regulatory bodies have approved drugs based on comparison with the previous SOC and clinical practice has established the path.

Cancer may be an acute disease if cured early, but sadly it is often a chronic disease leading to death. The medical goal is typically to begin with the therapy most likely to lead to cure. IO has unique ability to cure, but does so only in the minority of cases. Historically, trials that compared the average benefit of IO versus SOC that determine which is “better” have been the basis of approval. However, they do not address the most critical medical question of whether a patient’s path should begin with IO to deliver the best long-term outcome. This paper models the ability of “SMARTer” design to answer this question.

In some situations, IO may have a greater chance of curing, but SOC may on average create as many or more responses, creating a dilemma for patients, physicians and third-party payers. The modeling of SMARTer design presented herein, shows that even when IO has no greater benefit than SOC, if it is used first and patients transferred to SOC based on a rapid biochemical analysis of the effectiveness of the IO (an “optimizing test”), the total benefit may be better. We propose that this trial design would generate data suitable for the approval of that treatment path.

Approval of a treatment path is not novel. Use of the companion diagnostic has established the role of a test in defining a treatment path. Those positive by the test get one therapy, those negative by the test another. The “optimizing diagnostic” used herein is completely analogous to a companion diagnostic, however it is more accurate since it truly measures a patient’s initial response, while companion diagnostic attempts to predict if a patient will respond. Like a companion diagnostic, an optimizing diagnostic need not be 100% accurate to be useful. We modeled various sensitivities and specificities ranging from a “good” test with 95% sensitivity and specificity to “poor” 75% sensitivity and specificity. Tests measuring effectiveness are typically more accurate than tests attempting to predict response.

A “companion diagnostic” informs the choice of IO vs. SOC. An “optimizing diagnostic” informs the choice to remain on IO or transfer to SOC. In some trials intensification of IO based on a cfDNA optimizing test, or switching of one thymidine kinase inhibitor (TKI) to another TKI (based on detection of a mutation that causes resistance to the original)^14^ has been done. However, we are not aware of use of an optimizing diagnostic to change to SOC, within the experimental arm, or define a group for a second/sequential randomization to SOC alone or SOC with continuing IO.

A simultaneous combination of SOC & IO is often used for naïve patients as it is often better than either SOC or IO alone. However, direct comparison to sequential therapy is rarely available. Further, in the above example, the transfer of a significant sized group from IO (often for about 66% of the patients a test will determine continuing IO is not their best option) presents the opportunity or a sequential randomization to [SOC] vs. [SOC plus IO].

We first modeled a situation in which IO was better than standard of care showing a 40% response compared to a 30% response for SOC. Power analysis, for a standard IO vs SOC trial, using typical parameters, indicated that a trial of 710 people (355 each arm) would be needed to show that benefit. In contrast, assuming that the SOC was neither more nor less effective due to being delayed by the 3-6 week initial IO evaluation, a good optimizing test reduced the number of patients required to 114 (57 each arm).

In addition to requiring fewer patients and less time, more of these patients would receive the optimal therapy for them, either IO alone if that was sufficient, or IO and transfer to SOC, if that was best for the person. Thus, in addition to answering the key medical question of should a patient start with IO, the new design does so much more cost-effectively and meets a higher ethical standard. It facilitates trial completion in a timely manner both by cutting the number of patients required and enhancing accrual as patients are informed that if the new therapy is not working for them, they will be transferred to best SOC (or SOC will be added).

The presented modeling also shows that in the situation where IO and SOC are equal in a head-to-head trial. If initial IO is given with a near term optimization test with good performance, the path of IO followed by SOC would lead to a 63% improvement compared to SOC, assuming that IO neither potentiated nor inhibited subsequent SOC. Even when subsequent SOC was assumed to be 33% less effective, a good test and rapid switching resulted in the IO first path having a 40% benefit over SOC, despite them being equal in a head-to-head trial.

The literature is unclear concerning how delaying SOC would alter its benefit, thus we modeled multiple scenarios without favoring one. In clinical practice therapy often does not begin until 3 to 6 weeks after diagnosis; while delay may be assumed to disadvantageous, the magnitude of the change is unknown. To be conservative we modeled a 30% decrease in effectiveness as well as no change in effectiveness of SOC. Use of an optimizing diagnostic and SMARTer design is beneficial in both cases. We also modeled an enhancement of SOC as there is a growing body of evidence that initial IO sets the stage for better response to SOC, even if the IO itself fails. However the literature on this is complex and contradictory concerning if IO potentiates SOC. Maeda, et al., in 2019^15^ reviewed the general ineffectiveness of chemo for metastatic melanoma, with under 10% response rate and 1-2% durable responses, as well as multiple studies showing IO therapy does not decrease, and appears to increase, response rate to subsequent chemo; they reported unexpected good results with chemo after IO even in especially difficult to treat melanoma types. Also, in metastatic melanoma not expected to respond to chemo, Goldinger^16^ reported temozolomide after IO had a 3% response rate, consistent with no potentiation, but taxanes strikingly had a 27% response rate. Sato et al.^17^ reported IO potentiated taxanes in head and neck cancer. Schvartsman et al.^18^ in 2017 reported ORR was higher after failed IO than before. Park et al., in 2019^19^ found that last line salvage chemo after IO was more effective than the previous chemo before IO in NSCLC, while the expectation was that being 2 lines later it would be very much less effective. Mangin, in 2021^20^ noted IO appeared to enhance response to chemotherapy and reviews some studies that did not show this, concluding that it may depend on the subsequent chemotherapy type, consistent with Goldinger’s findings. Karachaliou et al., in 2021^21^, found chemo unexpectedly caused durable responses after failed immunotherapy. Puza^22^ found unexpected good results of surgery after IO. Saint-Jean^23^ reported potentiation of chemo following IO. Hadas-Bengad et al. in 2020^24^ reported IO potentiates chemo as did Markovic, et al.^25^ Asasa, et al.^26^, reported IO re-established sensitivity to cetuximab. While the literature is mixed and limited due to the retrospective nature of the studies, a body of studies suggests that in at least some cases IO potentiates subsequent chemo. Herein we propose prospective studies to evaluate the optimal path. While simultaneous combination has become standard for NSCLC, at least certain diseases with certain therapies, it may be less beneficial than the optimal sequential path. Kwong^27^ in preclinical work documented that chemo given at the same time as IO inhibited it, thus, the exact timing of IO is likely dependent on the specific chemo, specific IO, and specific disease. As with all oncology investigations, prospective multiple randomized trials, in this case of sequences, are needed and we propose should be included in trials designed for registration.

Theories as to why potentiation may occur include that while activating the immune system alone is insufficient for a response, that, in combination with subsequent killing (potentially with antigen release resulting in further immuno-stimulation) results in unexpectedly good response to subsequent SOC, in the setting of the IO altered immune system. Another possibility is IO may increase growth (PD or HPD) in a manner that makes cells more susceptible to SOC. The proposed sequential randomization design may provide additional data relevant to the question of whether IO increases the benefit of SOC more if given first or if continued with SOC, albeit only in the subgroup of patients that do not have a good biochemical response to IO alone.

The same methodology proposed herein can also be used to determine if treatment path(s) initiated with SOC can be optimized, as we presume they can. Jones, et al. presented an observational study (historical, not randomized) in an ASCO poster 2021, that observed no decrease in effectiveness of SOC if it was given after IO, but observed a potentiation of IO by SOC. However, no change in overall survival was seen. Zengin et al., 2021^28^ discusses, based on a failed study of simultaneous combination therapy, that sequential is potentially better, in this case chemo before immuno. Their discussion illustrates that even a statistically significant increase in PFS with combined therapy vs chemo alone was not enough to change clinical practice, emphasizing the importance of seeing larger group differences with methods such as those we propose. Interpretation of such analysis of historical, non-randomized data, is limited. The proposed sequential multiple randomized trial (SMART) procedure proposed herein can be modified to provide hard data as to the best sequence of therapies. “Reinforced Learning”^29^, “Deep Reinforcement learning”^30^, “Q-learning”^31^ and a “jackknife estimator”^32^ have been described as methods to optimize therapy and sequence.

While we used the example of IO, the proposed methodology applies to any new test.

Not surprisingly, models assuming potentiation of SOC by IO (from 30% to 40%) plus an optimizing diagnostic showed benefit over SOC, even when in a head-to-head trial of SOC vs IO, no difference would be seen. Surprisingly, the improvement over SOC was a dramatic - 86% with a good test. Even a poor test showed 47% improvement.

In addition to a design incorporating just use of an optimizing test, the use of sequential randomization to determine if SOC, or SOC plus IO, was more beneficial after switching, (in the subgroup with less likely to respond to IO) is also described herein. The result of the calculation of the benefit of the IO first group, assuming all patients received what was determined as the optimum second therapy, can be compared with SOC. Approval of the optimal path would be sought.

This study’s limitations include being a modeling study, that proposes a new type of trial, but does not present patient data.

Another limitation is that the performance of SOC in a subgroup that fails IO is not known, and may be outside of the situations modeled. (Our code or our collaboration is available for those wishing to model other scenarios.)

Another limitation is the performance of cfDNA testing, or other optimizing diagnostics, for progression is not established. Numerous reports correlate ctDNA levels with prognosis; more importantly, increasing ctDNA almost invariably predicts progression following immunotherapy, for instance Davis, et al.^33^, Bratman et al.^34^, and Herbreteau et al.^35^, each noted every patient with increasing cfDNA progressed. In fact, the latter and Wang et al.^36^, noted cfDNA testing had an AUC of 1, perfectly predicting response and detecting it, on average, 79 days earlier. However, as cancer therapies typically succeed for a period of time and then stop working, it is difficult to measure the accuracy of the cfDNA test for success of a therapy. It may be hundred percent accurate at showing that at a certain moment the therapy is working, but since the “gold standard” imaging is insensitive to changes at that time, and is typically only measured later, after the therapy may have stopped working, classic measures to evaluate test performance (a gold standard done at the same point in time) are not available. Thus, while we used a theoretical specificity, calculation of the actual specificity for progression is problematic. We modeled sensitivities of 95% and down to 70% and this may be highly conservative given the above references; our modeling of specificities may also be conservative and difficult to validate. However, different types of cfDNA testing may have different performance characteristics, the models apply to any optimizing diagnostic test.

In implementing a SMARTer design, a decision concerning the cut off for switching to the SOC would need to be made. In order to give the IO the greatest chance of working, likely the predefined formulae for transfer would be if there is evidence of increase in cancer; biochemically stable disease would remain on the IO. However, other cutoff points may be chosen; for example all patients not showing biochemical improvement might be transferred.

At the time of writing SOC does not include an optimizing test; SOC change based on biochemistry, rather than on imaging may evolve. If so, this should be included in the SOC arm. This trial design is only appropriate for drugs that have a different MOA. The above discussion focuses on use of cfDNA as a sample optimizing test. It is equally applicable to any method that can sensitively monitor cancer amount; examples include cancer specific exosomes, or advanced imaging with tracers. The modeling and discussion herein is based on the endpoint of “events”; survival to a certain timepoint may be considered an event, however survival analysis modeling was not performed herein.

In conclusion, clinical trials that include rapid evaluation of the new therapy by biochemical methods, and transferring to SOC if biochemical evidence suggests the new therapy is failing (an optimizing test), allow a new path to potential approval of new therapies. This path is analogous to a companion diagnostic, both specify whether the proposed new therapy should be used. Companion diagnostics specify if it should be initiated, optimizing test/diagnostics specify if it should be continued. New therapies that would fail in a head-to-head comparison with the previous SOC may be documented to enhance the patient path and provide a better medical outcome, potentially leading to approval with this new trial design. In one likely scenario where the new therapy is 30% better than the SOC, a head-to-head study would require over five times as many patients as one including rapid evaluation and therapy switching based on an optimizing diagnostic. A second randomization is proposed to determine if the path should be a transfer to SOC or to SOC plus continuing the new therapy. Use of such designs decreases the cost of clinical trials, is more ethical, likely improves accrual, and better models actual clinical use.

## Data Availability

All data produced in the present study are available upon reasonable request to the authors

